# Language fMRI lateralization success and head motion in pediatric epilepsy patients with ADHD, and improvements based on fMRI task training

**DOI:** 10.64898/2026.06.08.26355225

**Authors:** Bonnie Alexander, Kathryn Santamaria, Sila Genc, Sarah Barton, Mike Kean, Alison Wray, Wirginia Maixner, Emma MacDonald-Laurs, Joseph Y.-M. Yang

## Abstract

**Introduction:** Language functional MRI (fMRI) is a valuable tool for presurgical planning in epilepsy. Functional MRI can be challenging in children, and head motion can compromise its utility. The candidacy of patients with ADHD for fMRI is sometimes queried regarding concerns about possible head motion. In 2020, we implemented an fMRI task training program, via telehealth and/or mock MRI. We aimed to determine whether training increased language lateralisation success and/or reduced head motion in all patients, and in those with ADHD. We also aimed to determine whether patients with ADHD exhibited more head motion during fMRI than those without ADHD.

**Methods:** We retrospectively identified 223 epilepsy (85%) and other neurosurgery patients, (241 scans including repeats) with language fMRI at Royal Children’s Hospital, Melbourne, Australia, 2016-2024. There were 24 individuals with ADHD listed in the Electronic Medical Record, five of whom had diagnoses of both ADHD and autism; and nine with autism. Language lateralisation success was determined by clinician description recorded as left/right/bilateral in the medical record. 99 patients were provided the training including fMRI task practise. Head motion was quantified by maximum Framewise Displacement (FD_max_; mm).

**Results:** ADHD was associated with lower language lateralisation success. Training was associated with greater language lateralisation success, across all patients, and in those with ADHD. Regarding ADHD and head motion, outliers in FD_max_ were seen in 5 young patients with ADHD. Data were trimmed to allow separate investigation of FD_max_ for the sample with and without extremes of head motion. In untrimmed data, FD_max_ was significantly higher in patients with ADHD than in those without. In trimmed data, FD_max_ was on average lower in patients with ADHD than those without, however this was not statistically supported. Regarding training and head motion, across all patients, FD_max_ was significantly lower for scans with training than without. In patients with ADHD, FD_max_ was on average lower for scans with training, however training was not associated with FD_max_.

**Conclusions:** Language fMRI training was associated with higher language lateralization success, particularly in patients with ADHD. Training was associated with reduced head motion across all patients. Although some young patients with ADHD had substantial head motion, most in our sample did not move more than those without ADHD. We conclude that the training program increases success of language fMRI, and that an ADHD diagnosis should not be a contraindication to language fMRI.

## Introduction

Drug resistant focal epilepsy can be effectively treated with surgery (Harris et al., 2022; Jobst & Cascino, 2015; Macdonald-Laurs et al., 2021). Earlier surgical intervention is recommended particularly in children, as reducing the burden of seizures and anti seizure medication side effects on the developing brain can result in improved developmental outcomes (Braun, 2020; Cross et al., 2022; Holthausen et al., 2013; Jehi et al., 2022; Lamberink et al., 2020).

In determining surgical candidacy, many factors are considered (E.g., Ducis et al., 2016; Duncan et al., 2016; Jayakar et al., 2014) including language lateralisation and localisation which inform surgical risk of language impairment, and surgical boundaries (Benjamin, Li, et al., 2018). fMRI is favourable to the Wada test or surface electrode mapping for language mapping, as it is non-invasive and generally well tolerated (Benjamin et al., 2017; E.g., Liégeois et al., 2006). In children, feasibility of fMRI can depend on age, language abilities, behavioural challenges, and previous negative experiences with MRI scans and medical procedures (Cahoon & Davison, 2014). These factors may be particularly relevant in epilepsy, as children with epilepsy have increased prevalence of language difficulties (Parkinson, 2002), ADHD and autism spectrum disorder (which are also highly comorbid (E.g., Rau et al., 2020)), and other neurodevelopmental and neuropsychiatric conditions (Dagar & Falcone, 2020). The requirement to lie still for the 25-60 minute awake scan can be challenging for children, with excessive movement a potential contraindication to proceeding with the scan. Prolonged scan time can increase anxiety and/or movement, which may result in the scan being aborted (Hallowell et al., 2008). This may lead to a negative experience for the child and impact future medical interventions and procedures (Cahoon & Davison, 2014)

The suitability of patients with ADHD for presurgical fMRI is sometimes queried regarding concerns about possible excessive head motion. We have not identified any existing studies in clinical epilepsy presurgical fMRI quantifying head motion for patients with comorbid ADHD. There is however neuroimaging literature utilising standardised research fMRI acquisitions not acquired for clinical purposes, showing nuanced findings regarding success rate and head motion during fMRI for children with ADHD. Yerys et al. (2009) reported lower fMRI completion rates for 52 children with ADHD vs. controls, and lower success rate with younger age in an ADHD subgroup. In 56 9-11 year olds with ADHD and 61 controls, Thompson et al. (2021) report an association between ADHD and infrequent large head motion during resting state fMRI, mediated by scores on a sustained attention scale. Pardoe et al. (2016) noted greater head motion during resting-state fMRI in 332 ADHD vs. 549 controls in the ADHD-200 (Consortium, 2012) dataset, and greater head motion with younger age. Kong et al. (2014) reported the same finding in the ADHD-200 dataset, though a mediation analysis indicated that the larger head motion for ADHD group was largely attributed to individual differences in the behavioural measure of impulsivity. In a preregistered study, Shi et al. (2025) used an initial sample (624 ADHD of 971 total) and confirmation sample (296 ADHD of 437) from the Healthy Brain Network dataset (Alexander et al., 2017), reporting no associations between ADHD or ‘externalising disorders’ and head motion, but an interaction between age and neurodevelopmental disorders. The authors note that broad diagnostic categories may not be sufficient to understand head motion during fMRI, and suggest a more nuanced symptom-based approach incorporating caregiver report of children’s capacities may have more utility. By definition, ADHD is heterogeneous in presentation: The inattentive subtype is not characterised by excess motor activity, and in the hyperactive and combined subtypes, motoric hyperactivity decreases with adolescence (American Psychiatric Association, 2022). Further, a common reported feature of ADHD and autism is hyperfocus – highly sustained attention for fun or interesting tasks (E.g., Ashinoff & Abu-Akel, 2021). In the above studies, the distribution of head motion during fMRI is incompletely depicted, displayed as group means and summary statistics. Presenting individual data points would more accurately illustrate the range of head motion values In these individuals.

Providing patients and families with support and information can be facilitated in ways tailored to patients’ needs, including Mock MRI, and telehealth-based training. Mock MRI scanners are widely used to support patients through training and familiarization, while evaluating a child’s likely ability to lie still for the scan duration (Carter et al., 2010; Davis et al., 2022; De Bie et al., 2010; Gao et al., 2023; Wood et al., 2004). Mock scan use results in a greater proportion of successfully completed awake MRI scans (De Bie et al., 2010). In non-medical research settings, mock scanning has been used to assist neurodivergent children with fMRI. Epstein et al. (2007) administered head motion training during Mock fMRI, finding no difference in success rate of cognitive task fMRI between ADHD and controls. Pua et al. (2020) prepared children with autism for MRI, including Mock MRI training, reporting similar or better fMRI image quality metrics for to individuals with autism in the ABIDE II (Di Martino et al., 2017) cohort.

Telehealth, i.e., health appointment via video call, is widely used, with benefits to families including access to resources, education, training, and support (Kissani et al., 2020; Macwilliam et al., 2021). It is an accessible means to build rapport and to provide training in the specific tasks required for language fMRI. Karakas et al. (2015) provided task fMRI training and scan preparation to 6-12 year old boys with ADHD, demonstrating no difference in acquisition success rate between ADHD and controls. However, there was no condition without training, so the effect of training could not be explicitly tested. Language tasks may be difficult or infeasible in children with language limitations (De Guibert et al., 2010). In a presurgical evaluation context, supporting children with pre-scan training in performing the specific functional tasks allows for time for familiarisation; and also the potential to delay scan to allow for development, or discontinue if not appropriate.

In 2020, we implemented a functional MRI training program using telehealth for children being considered for an fMRI scan for language lateralisation at the Royal Children’s Hospital (RCH), Melbourne. Where indicated, Mock MRI was incorporated into the pre scan training. Children were triaged by the Epilepsy Nurse Consultant and then proceeded to formal fMRI training using telehealth and/or mock MRI. The current project is a retrospective analysis in epilepsy and neuro-oncology patients at RCH, between 2016-2024, who were ordered language fMRI scans for presurgical evaluation. We wanted to evaluate the effectiveness of this program for this cohort, including specifically for those with ADHD. Although autism spectrum disorder is frequently comorbid with both ADHD and epilepsy, it was outside of the scope and statistical power of the current paper investigate as an additional factor, however it is included descriptively to complement our central clinically-driven question about ADHD. We aimed to determine whether pre-scan training increased language lateralisation success and/or reduced head motion in all epilepsy and neuro-oncology patients, and in those with ADHD. We also aimed to determine whether epilepsy and neuro-oncology patients with ADHD had less successful language lateralisation, or more head motion during language fMRI, than those without ADHD.

## Methods

The Human Research Ethics Committee of our hospital approved this retrospective study (HREC 36328), and the requirement for informed consent was waived due to the retrospective nature of the investigation.

### Sample selection

We retrospectively identified epilepsy and other neurosurgery patients with language fMRI at the RCH, 2016-2024. Data were gathered by screening all instances of the item code ‘MRI brain functional’ since 2016 when the electronic medical record (EMR) was introduced. These instances were refined based on language fMRI task data acquired as reported in the EMR and data listed on either Synapse or PACS. Exclusion criteria were: age > 18 years, language fMRI not clinically processed due to patient deceased, major structural brain abnormality such as missing lobe or hemisphere. While the language lateralisation variable was determined for the whole sample, a subset of the sample had data collected for head motion, due to constraints on data reprocessing described below.

### Demographic and clinical data collection

Demographic and clinical information were collected from the EMR. ADHD and/or Autism were derived from plain text descriptions in the EMR noting existing diagnoses. For patients with ADHD, subtypes of ADHD, and ADHD medication status, were not always listed in the EMR; and no data existed on whether ADHD medication was taken on the day of scan. Thus, these factors were not investigated.

Data on provision of the training program via telehealth was stored in departmental databases.

Because the time span of the sample extended to before Mock MRI sessions were documented in EMR, it was not possible to completely quantify how many patients had a mock and therefore to test the effect of Mock MRI for the current sample. However, approximate numbers were gathered.

### Outcome measure data collection

Language lateralisation success was a binary categorical factor with success indicated by clinician description of language fMRI laterality as left/right/bilateral in the EMR. Non-success was attributed to scenarios including uninterpretable data, such as absence of language network activation; or incomplete data, such as scan discontinued due to patient discomfort. One patient’s data was classified as not successfully lateralised due to absence of laterality described in EMR, but was reclassified to successfully lateralised after reprocessing with fMRIPREP as described in below sections and in Supplementary Information.

Head motion during language fMRI was estimated using Framewise Displacement (described further in methods), based on standardised fMRI preprocessing with fMRIPREP, which was utilised clinically since 2020 and run on older data for the current project. For some scans, fMRIPREP did not complete. Clinically, other accepted methods had been used in processing some data. In these cases, head motion was not recorded.

### Language fMRI task training procedure

Patients at the RCH under the care of the neurology or neurosurgery departments who were ordered a language fMRI scan, were triaged through the training program as illustrated in Figure 1.

**Figure 1.**
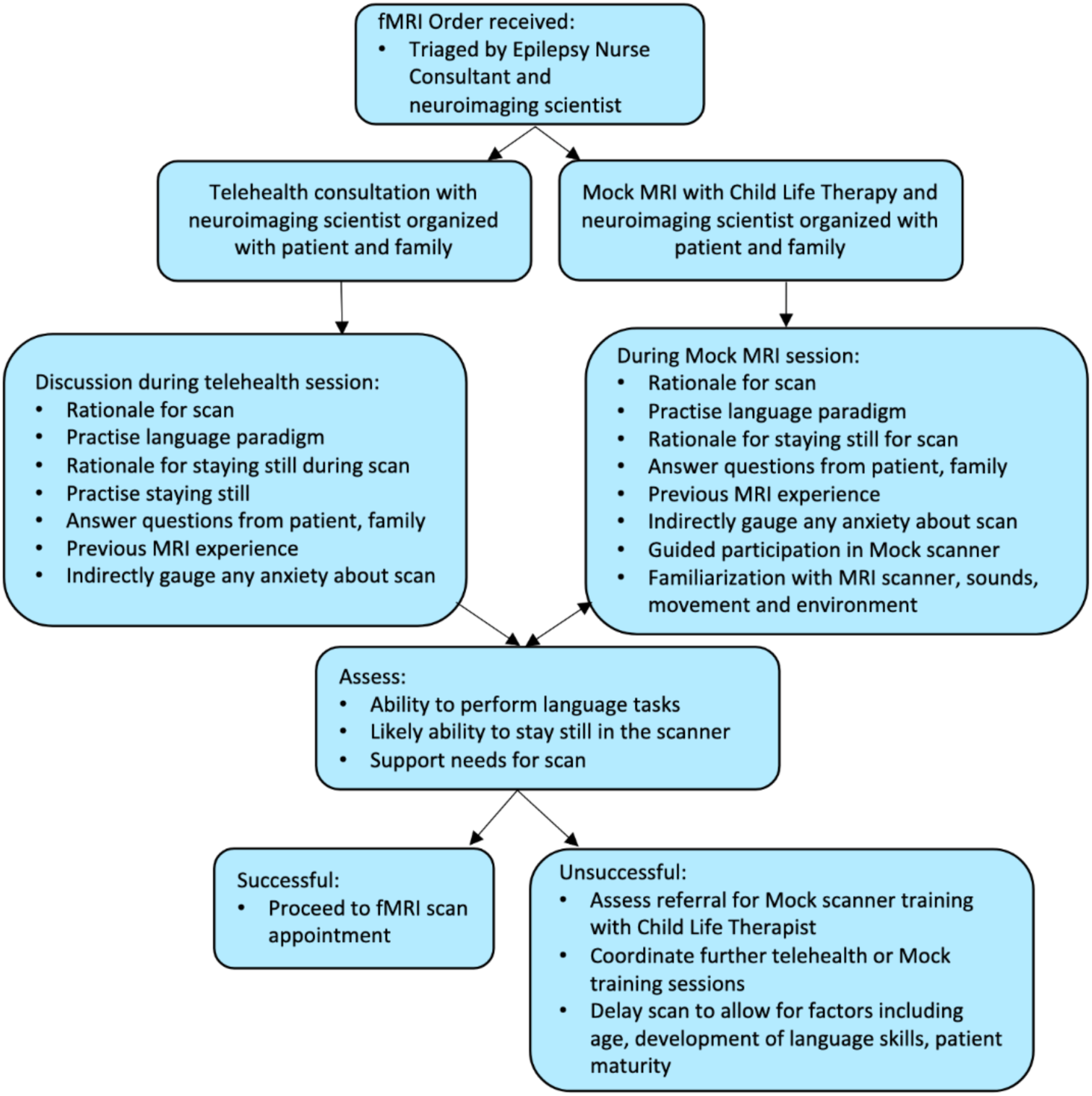
Language fMRI training program workflow. An example of the dialogue with the patient and family provided during task training via telehealth is included in Supplementary Material.

Training (either via telehealth or in-person Mock MRI) included patients practising the language tasks that they would need to complete during the fMRI scan. This included a Verb Generation and an Orthographic-Lexical Retrieval task as described in Wood et al. (2004). These tasks require individuals to generate nouns and verbs directly correlating to a sequence of visual prompts.

### MRI acquisition and processing

MRI was acquired on a 3T Siemens scanner at Royal Children’s Hospital, Melbourne, Australia (Tim Trio 2016-September 2017, Prisma Fit September 2017 onwards).

### Language task

The language task was a Verb Generation paradigm (Wood et al., 2004) where patients were visually presented with nouns (such as ‘dog’) and were asked to imagine a corresponding verb (such as ‘bark’) in their mind without speaking or mouthing the words. This task was a block design with 36-second alternating blocks of four rest and three active conditions. In the rest condition, a crosshair was presented; and in the active condition one stimulus was presented either every 4 seconds, or every 8 or 9 seconds, based on patient needs determined by the neuroimaging scientist with the patient and family during task training. Nouns presented are those listed in Wood et al., or short, phonetically spelled nouns like ‘cat’ ‘sun’ ‘ant’ for patients unable to read longer words.

### MRI sequences

T1 images were acquired with 0.8 mm isotropic voxels, optionally zero-filled interpolated to 0.4 x 0.4 mm in-plane. Language task Echo Planar Imaging data was acquired with either Siemens product or CMRR 2D EPI sequences. Siemens sequences had: 38 slices, slice thickness 3.15 mm, 1.88 x 1.88 mm voxels in plane, FOV 240 x 240 mm, TR 3 s, TE 40 ms, flip angle 90, multiband factor 2 with GRAPPA, with GRE field maps for magnetic susceptibility distortion correction. CMRR sequences had: 60 slices, slice thickness 2.5 mm, in plane resolution 2.45 x 2.45 mm, FOV 255 x 255, TR 1.5 s, TE 30 ms, flip angle 70, multiband factor 3 with GRAPPA, and Blip Up and Blip Down pairs for distortion correction.

### Preprocessing and processing

For routine clinical processing since April 2020, raw DICOM data were converted to NIFTI in BIDS (Gorgolewski et al., 2016) structure using ‘dcm2bids’ v2.1.6 (Bore et al., 2023). fMRI was preprocessed with fMRIPREP (Esteban et al., 2019) including motion correction, slice timing correction, susceptibility distortion correction, and registration to T1, output in patient space; and was processed using FEAT (Jenkinson et al., 2012). Data from 2016 to April 2020 were previously not preprocessed with fMRIPREP, and were reprocessed with fMRIPREP and FEAT for standardised preprocessing and estimation of head motion. Some fMRIPREP runs did not complete, described in results. For reprocessed data with newly completed fMRIPREP and FEAT runs, language task outputs were inspected (by authors K.S. and B.A.) to ascertain whether language laterality could be determined and confirm laterality matched with that described in the medical record. The results of this process are described in Supplementary Information.

Head motion was quantified using Framewise Displacement (FD; mm), calculated by the summed absolute values of change in six motion parameters (cumulative translation in x, y and z, and cumulative pitch, yaw and roll) for each volume (Power et al., 2012). fMRIPREP outputs FD for each volume, however, this automated metric is confounded by sample rate in our clinical data comprising older sequences with TR = 3 and newer sequences with TR = 1.5. To address this confound, we estimated FD based on every second volume of the CMRR sequences, termed ‘harmonised’ FD.

### Statistical analysis

To investigate associations between language fMRI training, ADHD, and the outcome variable of language lateralisation success, Generalised Estimating Equations (GEE) models with binomial distribution and logit link function were employed. Age at scan and sex were also included in the model, however were not the primary factors of interest. Repeated measurements were accounted for by clustering on patient ID. The model was run using the ‘geeglm’ function in ‘geepack’ package in R (R Core Team, 2023). This function handles repeat observations and was also robust to perfect separation seen in the current data where all observations of repeat fMRI with training had the same value on the outcome variable. The following model was used to (test) effects of ADHD, and training, on language lateralisation success across the whole sample: *gee_all_patients = geeglm(lang_laterality_determined ∼ training_program + age_at_scan + adhd_binary + sex, family = binomial(link=“logit”), id = patient_id, corstr = “exchangeable”, data = th_df).* To test the effect of training in patients with ADHD, the following model was used: *gee_in_patients_with_adhd = geeglm(lang_laterality_determined ∼ training_program, family = binomial(link=”logit”), id = patient_id, corstr = “exchangeable”, data = th_df_adhd_subset)*.

To investigate associations between language fMRI training, diagnosis of ADHD, and FDmax, linear mixed effects models were used using ’lmer’ in package ‘lme4’ (Bates et al., 2015) in R. The following model was used to test the effect of ADHD and training on harmonised FD_max_ across all patients: *lmer_harmonised_max_fd <- lmer(harmonised_max_fd ∼ training_program + age_at_scan + adhd_binary + (1 | patient_id), data = th_df).* To test the effect of training on FD_max_ in patients with adhd, the following model was used: *lmer_harmonised_max_fd_in_patients_with_adhd <-lmer(nv_maxfd_harmonised ∼ training_program + age_at_scan + (1 | patient_id), data = th_df_adhd_subset)*.

## Results

### Sample

We retrospectively identified and screened 296 acquisitions labelled ‘MRI brain functional’ in EMR within the time range 1st January 2016 – 17^th^ April 2024. 55 acquisitions were excluded for the following reasons. 13 acquisitions did not include language tasks (13 motor fMRI only, 24 resting state fMRI only, 1 EEG-fMRI, 1 memory fMRI, 1 not determined, 1 DWI only), 2 duplicate records, 3 data not processed at RCH, 1 data not processed due to patient deceased, 2 hemisphere/lobe not present, 6 over 18 years old.

The resultant sample comprised 223 epilepsy and other neurosurgery patients (241 scans including repeats) with language fMRI at Royal Children’s Hospital, 2016-2024 (age at scan 6.04-17.93 years, *M* = 12.75, *SD* = 2.89, Mdn = 12.73; 111 scans female/ 130 scans male). Departmental databases since April 2020 indicated request pathways through epilepsy program (85%) and neurosurgery (15% including neurooncology and neurovascular cases some of whom also had seizures). There were 24 individuals with ADHD listed in EMR, five of whom had both ADHD and autism; and nine with autism. There were two patients noted to have suspected but not recorded autism, one of whom had ADHD, and one with no ADHD. Demographic information summarised by diagnosis of ADHD and/or Autism is included in Table 1.

**Table 1.** Total sample demographic information summarised by the categorical factors of ADHD and/or Autism.

| ADHD and/or Autism | n individuals | Age at earliest scan in dataset<br>Min-max (mean (SD)) | Age across scans (including repeats)<br>Min-max (mean (SD)) | Sex (female/male) | Training program provided for scan (incl repeats) (n) |
| --- | --- | --- | --- | --- | --- |
| ADHD*# | 19 | 6.59-17.9 (10.9 (2.88)) | 6.59-17.9 (10.9(2.69)) | 7/12 | 11 |
| Autism*# | 9 | 9.04-14.9 (12.1 (2.29)) | 9.04-14.9 (12.1(2.29)) | 4/5 | 0 |
| ADHD+autism*# | 5 | 9.39-12.6 (10.8 (1.48)) | 9.39-13.3 (11.3(1.67)) | 1/4 | 3 |
| No ADHD or Autism (total)* | 190 | 6.04-17.8 (13.0 (2.87)) | 6.04-17.8 (13.0, 2.88) | 90/100 | 87 |
| No ADHD or Autism (with complete acquisition and processing)# | 173 | 6.86-17.8 (13.0 (2.79)) | 6.86-17.8 (13.1, 2.77) | 85/88 | 87 |

\*Indicates demographic summary relevant to comparisons of language lateralisation success
#Indicates demographic summary relevant to comparisons of head motion during fMRI

**Table 2.** Total Sample demographic information summarised by categorical factor of training program provided or not provided.

|  | n scans | Age at scan<br>Min-max (mean (SD)) | Sex (female/male) |
| --- | --- | --- | --- |
| Training program | 101 | 7.29-17.8 (12.6 (2.61)) | 40/61 |
| No training program | 140 | 6.04-17.9 (12.9 (3.08)) | 71/69 |

The subsample with complete fMRIPREP and FEAT runs, i.e., used in comparisons of head motion during fMRI, comprised 208 patients (total 223 scans including repeats; age at scan 6.59 – 17.93 years, *M* = 12.77, *SD* = 2.81, *Mdn* = 12.73, 105 scans female, 118 scans male). The reasons for incomplete data acquisition or processing are as follows. For 4 patients, scan acquisition was discontinued due to patient discomfort. For one patient there was excessive artifact from dental braces. For 12 patients, data acquired prior to April 2020 (i.e., not preprocessed with fMRIPREP) could not be located or successfully retrieved. For 1 patient, fMRIPREP did not handle specification of field maps. All instances of incomplete acquisition or processing were in individuals with no diagnoses of ADHD or autism. Demographics for the subset with complete fMRIPREP and FEAT data are included in a separate row in Table 1.

Ninety-nine individual patients (101 scans) were provided the fMRI training program. Wilcoxon rank sum test indicated no evidence for difference in age between scans with training provided and scans with no training provided (*W* = 7776, *p =* .3).

In person mock MRI training prior to April 2020 comprised 3 female patients, 2 of whom had known ADHD, and none had autism. In person mock MRI training post April 2020 comprised 5 female and 10 male patients. 4 of these patients had known ADHD, 1 patient with suspected autism, 2 patients with ADHD + autism and 10 patients with no ADHD or autism.

### Training program

Since April 2020, 99 of 114 patients (101 of 117 scans) who had language fMRI were provided the training program.

Instances where training was not provided comprised 3 urgent scans, 2 no staff available, 2 patients did not attend, 4 phone call and/or training in scanner room prior to scanning instead, 1 abridged training provided during acute clinical deterioration prior to urgent scan, 4 not documented.

Of the cases where the training program was utilised, the triage process as in Figure 1 was followed to provide support according to patient needs. It was determined by nurse triage (reflective of nursing clinical expertise and experience with fMRI candidates) that 2 did not need telehealth or mock and proceeded directly to scan, 2 patients received phone calls that determined training was not needed, 76 were provided telehealth training, roughly 7 provided mock, 14 had in-person training in outpatient or inpatient clinical setting. For simplicity of terminology and analysis in the current paper, all these instances are subsequently referred to as “provided the training program” as they have been triaged through the process documented in Figure 1.

### Language lateralisation success

There were 33 scans with language laterality categorised as not determined (i.e., not listed as left, right, or bilateral in EMR). These data included the following scenarios. For five scans, the acquisition of the Verb Generation task was discontinued due to patient discomfort including three instances (two in one patient) of discomfort with scanner sounds, one claustrophobia, one complex circumstance including urgent scan in young patient with medication side effects causing discomfort. Twenty-eight scans had Verb Generation task data that was inconclusive. Where there were accompanying free-text notes in the EMR providing context for the inconclusive result, these included: Three notes of no activation for Verb Generation task, one note of activation in areas other than language network, six notes of motion artifact marring data, one note of artifact due to dental braces, one note of short sightedness, one note regarding poor verbal fluency impacting task performance, one note of hyperactivity and attention deficit, and one note speculating attention may have been on keeping still rather than performing language task.

ADHD and language lateralisation success

For patients with ADHD, 72.4% of scans had successful language lateralisation, compared with 88.6% in patients without ADHD. GEE model indicated ADHD was associated with lower language lateralisation success (*p* = .028, OR = 0.30, 95%CI 0.101-0.857).

Training and language lateralisation success - in all patients

Language lateralisation was successful in 112 (80.0%) of patients without training, and 97 (96.0%) patients with the fMRI telehealth training (Figure 2(a)). GEE model indicated a significant, strong association between the training program and successful language lateralisation (*p* < .001; OR 7.66, 95%CI 2.39-24.6). Age was not significantly associated with language lateralization success (*p* = .197; OR 1.10, 95%CI 0.95-1.27). This can be clearly appreciated in Figure 2a, across all ages of the sample: without training (circular points), unsuccessful scans can be seen across the age distribution.

**Figure 2.**
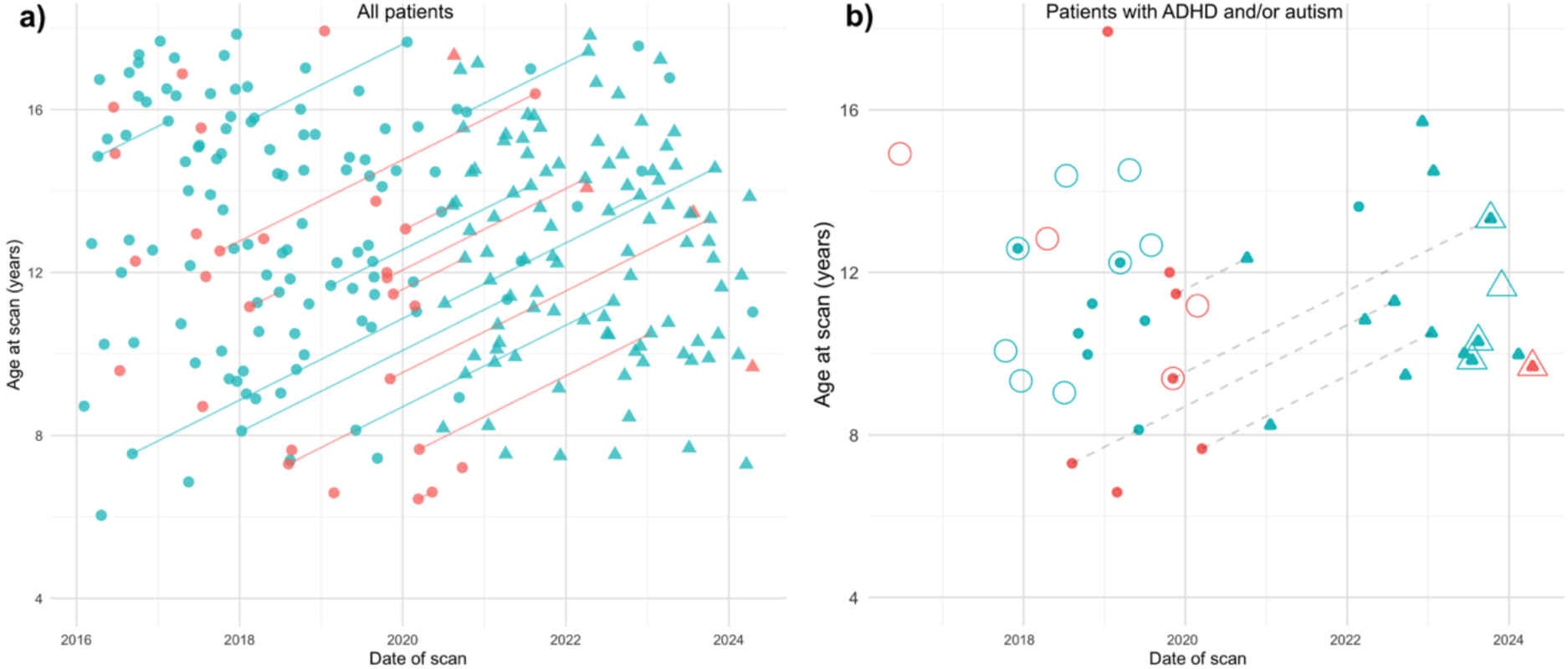
Success of language lateralization, fMRI training, and age. a) For all language fMRI acquisitions. b) for epilepsy patients with ADHD and/or Autism. Green points: language laterality determined, red: language laterality not determined. Triangles: fMRI training provided, circles: fMRI training not provided. Connecting lines: repeat scans for individual patients. For b): Small filled points: ADHD, large hollow points: autism.

Sex (male) was associated with less successful language lateralisation (*p* = .044; OR 0.43, 95%CI 0.20-0.98), the factor of training remained associated with language lateralization success independent of sex. It can also be appreciated that in all cases of repeat scans (indicated by points joined by a line) with training, language lateralisation was successful. This constitutes statistical perfect separation, precluding the factor of ‘repeat’ from being explicitly tested with this model. However, logistic regression in only first instances (*n* = 212) of language fMRI (detailed in Supplementary Material), indicated training was strongly associated with language lateralisation success, supporting that the effect of training could not be explained by repeat of language fMRI.

Training and language lateralisation success - in patients with ADHD

For patients with ADHD with no language fMRI training, 8 scans (53.3%) resulted in successful language lateralisation. For patients with ADHD with training, 13 scans (92.9%) resulted in successful language lateralisation (Figure 2(b)) GEE model indicated training was associated with language lateralisation success in patients with ADHD (*p* = .039, OR = 11.0, 95%CI 1.13-108). Age was not significantly associated with language lateralisation success (*p* = 0.544, OR = 1.16, 95%CI 0.72-1.85)

### Head motion during clinical language fMRI acquisition

Scatterplots of maximum and mean harmonised Framewise Displacement (FD_max_) during language task fMRI are shown in Figure 3, for patients with and without diagnoses of ADHD. For completeness, scatterplots using only CMRR sequences (which have TR = 1.5s) with original FD_max_, are included in Supplementary Material.

**Figure 3.**
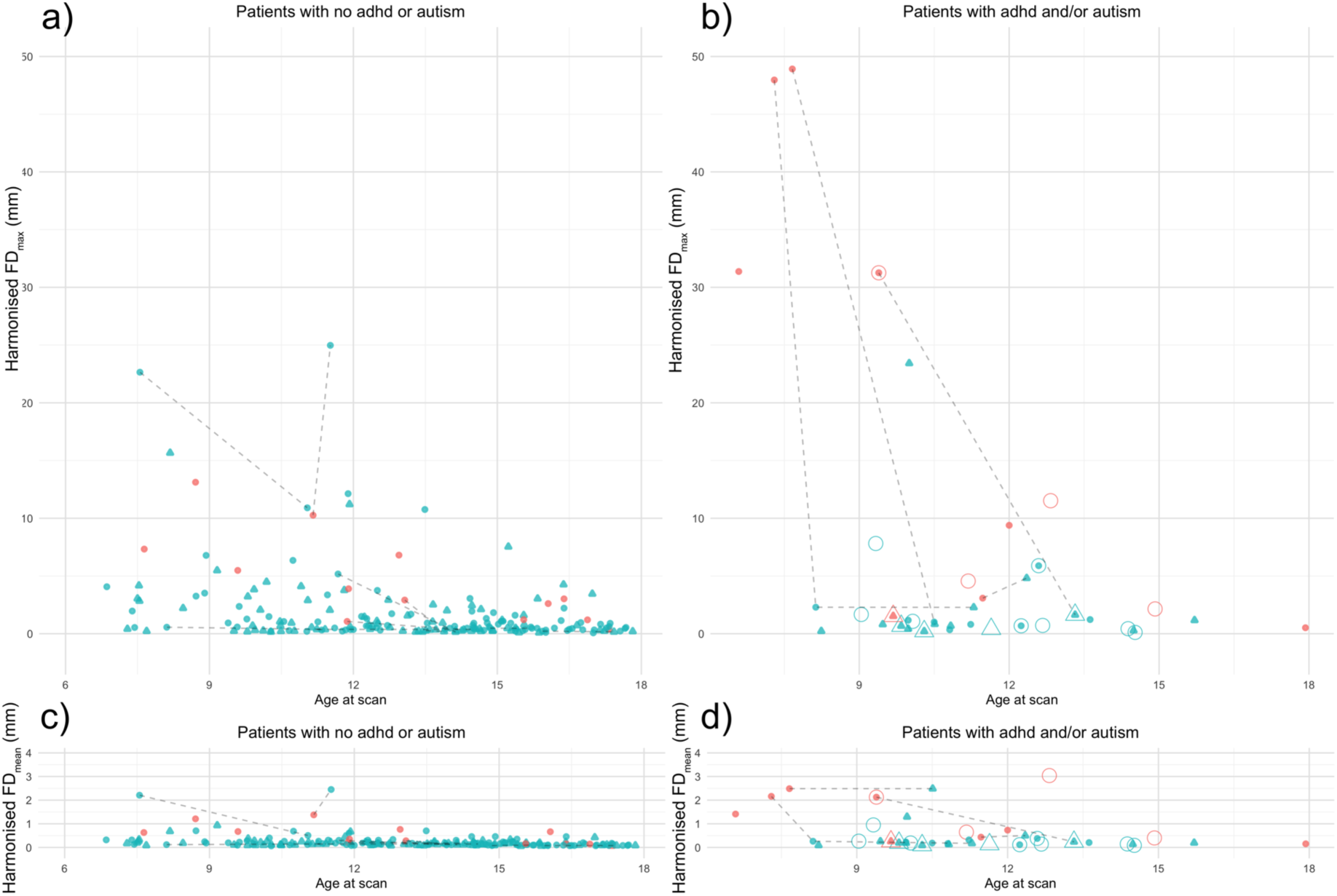
Head motion (harmonized Framewise Displacement in mm) during language fMRI, age, training, and success of language lateralization, for epilepsy patients with ADHD. A) and b): maximum harmonized FD, c) and d): mean harmonized FD. Green points: language laterality determined, red: language laterality not determined. Triangles: fMRI training provided, circles: fMRI training not provided. Connecting lines: repeat scans for individual patients. For b) and d): Small filled points: ADHD, large hollow points: autism.

ADHD and head motion

Extreme outliers in FD_max_ (see Fig.3 (a), (b)) were noted, where FD_max_ > 20mm. In patients with ADHD, these data visually appear to cluster in five patients < 11 years old. Data were trimmed to FD_max_ <20 to allow separate investigation of the rest of the distribution. In untrimmed data, in patients with ADHD, harmonised FD_max_ was on average higher (*M* = 7.76, *SD* = 14.2, Mdn = 1.17) than in patients without ADHD (*M* = 2.04, *SD* = 3.46, Mdn = 0.72). Linear mixed effects models indicated ADHD was associated with higher harmonised FD_max_ (*p* < .001, estimate 4.33, 95%CI 2.05-6.61). In trimmed data, in patients with ADHD, harmonised FD_max_ was on average lower (*M* = 1.79, *SD* = 2.20, Mdn = 1.01) than in patients without ADHD (*M* = 1.83, *SD* = 2.68, Mdn 0.723). However, linear mixed effects model did not indicate evidence for ADHD being associated with harmonised FD_max_ (*p* = 0.357, estimate 0.58, 95%CI .184-1.85)

Training and head motion - in all patients

In all patients, for scans with training, harmonised FD_max_ was on average lower (*M* = 1.71, *SD* = 3.16, Mdn = 0.63) than for scans with no training (*M* = 3.64, *SD* = 7.86, Mdn = 0.95). Linear mixed effects model indicated language fMRI training was associated with lower harmonised FD_max_ (*p* = .003, estimate −2.30, 95%CI −3.79 - -0.80).

Training and head motion - in patients with ADHD

In patients with ADHD, for scans with language fMRI training, harmonised FD_max_ was on average lower (*M* = 2.79, *SD* = 6.06, Mdn = 0.82) than for those without language fMRI training (*M* = 12.4, *SD* = 17.9, Mdn = 2.29). Training was not associated with harmonised FD_max_ (*p* = 0.063, estimate 0.01, 95%CI −0 – 1.6).

## Discussion

This study was a retrospective study utilising clinically acquired task-based language fMRI used for presurgical planning and evaluation in a sample of pediatric epilepsy and neuro-oncology patients, some of whom also had ADHD. We measured success of language lateralisation, and head motion during language fMRI. We found that ADHD was associated with these outcome measures, though with some nuanced patterns of results. We also found that the training program overall was associated with improved lateralisation success and decreased head motion during language fMRI, with details and limitations discussed below.

## Language lateralisation success

### ADHD and language lateralisation success

Across all patients with ADHD in the study, the success rate of language lateralisation was 72.4%, ∼16% lower than in those without ADHD. This figure comprises 53.3% success rate in those without training and 92.9% in those with training, as discussed further below. Regarding existing literature, there does not appear to be an equivalent sample and design in previous research on scan success rate, either in terms of comorbid epilepsy or neuro-oncology and ADHD, or in terms of use of clinical presurgical language fMRI. In comparison with studies in non-clinically acquired resting state fMRI, this finding re the association of ADHD with scan success rate is in the same direction with that from Yerys et al. (2009) who found lower fMRI battery completion rates (∼78% vs 96%) in participants with ADHD compared with those without ADHD. These authors also report lower battery completion rate (80% vs 84%) in participants with epilepsy vs controls, however their study does not contain a subsample with both conditions.

### Training and language lateralisation success

The training program was found to be strongly associated with a higher success rate across the whole sample, with ∼17% higher success rate and 8x higher odds of success with training. Age was not associated with language lateralisation success, as shown in some older patients with unsuccessful lateralisation both with and without ADHD. This is consistent with some previous findings re age and completion of fMRI, e.g., Yerys et al. (2009) reported that epilepsy and control participants 10 and above had higher fMRI battery completion rate than 4-6-year-old groups, and report no significant differences based on age for ADHD or ASD participants. In controls collapsed across studies, 10-12-year-olds had higher fMRI battery success rate than all other age groups. Particularly in a clinical setting, factors unrelated to young age might result in incomplete scans or unsuccessful language lateralisation, such as individual differences in cognitive functioning, claustrophobia in older individuals, other medical contextual factors including epilepsy. We expect that training would likely be useful in an adult epilepsy or neurosurgery presurgical planning setting.

In patients with comorbid ADHD, the training program was associated with a ∼40% higher language lateralisation success rate and 11.4x greater odds of success. There do not appear to be comparable studies explicitly testing the effect of fMRI task training in a clinical cohort with ADHD, so the current study newly contributes to the literature on this point.

We speculate the training was effective due to patients being informed about the purpose and procedure of the scan, and motivated and confident to perform the language task. Increased understanding and decreased uncertainty or anxiety about the task and any other situational factors may have lead to better cognitive performance of the task during scan, and thus better signal in the language network. However, task performance, anxiety or confidence before or during the scan was not directly measured, so we cannot draw further inference about the mechanisms underlying scan success. We expect that for some patients where head motion may otherwise preclude successful lateralisation, understanding and motivation to stay still may help, as discussed further below.

Another important point is that the triage process itself helps screen out patients who were unequivocally unable to perform the tasks due to head motion, attention or other neurodevelopmental factors, in turn likely leading to a higher success rate. As described in Figure 1, some patients’ scans may be delayed to allow for age and neurodevelopmental maturation where appropriate and feasible.

## Head motion during language fMRI

### ADHD and head motion

The association of ADHD with greater head motion is broadly consistent with some findings in the neuroimaging research field utilising resting state fMRI (Kong et al., 2014; Pardoe et al., 2016; Thomson et al., 2021), but not consistent with, e.g., Shi et al. (2025) who found no difference. Across the 24 patients with ADHD, harmonised FD_max_ (i.e., the maximum displacement in mm between 3-second-apart samples across the acquisition) during language fMRI was on average 5.7 mm greater than for those without ADHD. However the median FD_max_ was 0.35 mm greater. This difference in average FD_max_ was found to be due to the presence of extreme outliers, in five young patients with FD_max_ of 24 – 50 mm across 3 seconds. Figure 3b) illustrates that even in young patients with ADHD, most harmonised FD_max_ values are between 0 - 2 mm, with some around 0.2 - 0.5 mm. Figure 3d) shows FD_mean_, rather than FD_max_ across the task timecourse, for patients with ADHD, indicating many patients including 8-year olds who had average < 0.5 mm head motion between each volume across the task. Removing the extreme outliers resulted in patients with ADHD having a lower average harmonised FD_max_ for than those without ADHD, however this was not a statistically supported difference. Displaying all datapoints as per Figure 3 is critical to understanding the distribution of head motion for this cohort of patients. Although not bimodal, it is not normally distributed and summary statistics should not be interpreted as if per a normal distribution.

### Training and head motion

Language fMRI training was associated with lower harmonised FD_max_ across the sample, i.e., a median FD_max_ of 0.63 mm with training compared with 0.95 mm without training. This is expected in turn to improve data quality, and validity for both lateralization and localization for surgical planning.

In the 24 patients with ADHD, scans with training had a median harmonised FD_max_ of 0.82 mm, compared with median harmonised FD_max_ of 2.29 mm for scans without training. Though the statistical support for this was weaker, limiting generalisability, this is expected to have been beneficial in terms of data quality in our sample.

The presence of volumes with large framewise displacement is not always prohibitive of interpretability and lateralisation. In Figure 3 a) and b), there are some datapoints with FD_max_ between 20-25mm, with successful language lateralisation. Speculatively, this may relate to the robustness of the block design task vs. other task designs or resting state (e.g., Smith et al., 2013). Block designs are frequently used for clinical language fMRI (Benjamin, Dhingra, et al., 2018). We did not have an exclusion criterion of large FD in this study or in clinical processing, because despite noise that may compromise the precision of localisation re surgical boundaries, any valid information on laterality remains important. Another factor could be patients performing task very effectively, providing strong BOLD differences between task and rest.

### Limitations

Alongside the clinical applicability and substantial sample size afforded by this retrospective design, there are multiple limitations of the current study.

There are almost certainly more patients in the sample with ADHD or autism, who have not sought or received a diagnosis. Further, primary records of formal diagnostic reports of ADHD and autism would be a preferable source for these categorical variables rather than clinician notes of existing diagnoses formally documented outside of our hospital databases. The retrospective nature of this study means the extant data in EMR is the best information we have on this. Relatedly, understanding and diagnosis of ADHD, Autism and related conditions has increased greatly in recent years, so earlier data in the sample may disproportionately underrepresent patients with ADHD and/or Autism. Prospective studies may be better placed to more completely characterise patients with this subset of conditions.

In the current study, diagnosis of Autism was included in descriptive statistics and plots of data in response to queries regarding comorbidity, and how success and head motion are distributed in patients with Autism. However, Autism was not investigated or statistically tested as an additional predictor. Thus the datapoints are included in plots for completeness of illustration, and may serve as hypothesis-generating data for future work.

Subtypes of ADHD, ADHD medication status, and individual differences in cognitive factors such as memory, and attention, were also not all clinically investigated or recorded for each patient in the EMR. These factors were not investigated in the current paper, and we cannot draw inference on their possible influence on outcome variables. Limitations on subsample size would also likely have precluded statistical power to differentiate these factors. Prospective study designs would be better suited to answering questions about these factors and would be valuable to add nuance to understanding of how to best support patients based on their individual needs and strengths.

The presence of two different Echo Planar Imaging sequences in the dataset is a possible confounding factor for the measure of language lateralisation success and possibly head motion. The older (Siemens product) sequences have a smaller field of view and lower signal to noise ratio than CMRR sequences, however are less susceptible to motion artifact seen with additional multiband factor and GRAPPA. The CMRR sequences were adopted in 2018 (see Supplementary Material for figure illustrating sequence per scan), and the training program was introduced in 2020, so sequence partially confounds the factor of training. We believe using harmonised FD_max_ is the most effective method of minimising this confound on the head motion data, given the fortunate circumstance of a TR for CMRR sequences of exactly half of that for the Siemens sequences.

### Clinical value and recommendations

Overall, we have found the training program to be a valuable addition to our clinical assessment pathway, in the higher success rate and data quality (i.e., less head motion) of language fMRI associated with training. Successful acquisition of high quality fMRI data affords more accurate estimation of surgical risk and surgical boundaries.

There are health economic benefits to increased success rate. Spared fMRI slots avoid delayed investigations and delayed care for current and other patients. As the telehealth sessions are short (∼15 minutes) and straightforward, and can be conducted by, e.g., imaging scientists and epilepsy nurses, they do not necessitate an added cost burden.

The current project demonstrates the value of a patient centered approach in maximising the opportunity for patients with ADHD to complete this important investigation. Delivering information tailored to the individual’s capabilities may be assisted by understanding how to interpret neurodivergent patients’ behaviour outside of the scanner. For example, drifting gaze may indicate thinking (Doherty-Sneddon et al., 2009; Doherty-Sneddon et al., 2012; Martinez-Cedillo et al., 2025), and fidgeting or physically moving may indicate self-regulation (Aspiranti & Hulac, 2022; Tancredi & Abrahamson, 2024; Zentall & Meyer, 1987) rather than inattention. Speculatively, playing with toys or wriggling during conversation and instructions may serve as self-soothing for sensory and medical contextual factors, or overcoming boredom, understimulation, uncomfortable power dynamics, or other emotions. The same patients may stay very still during task examples and scanning, when understanding the rationale of taking a clear picture to help with medical investigations. Attention in ADHD is suggested to be highly focused based on interest or importance (Ashinoff & Abu-Akel, 2021), and it may be possible that concordantly, the clinical importance of the scan and/or interest in the process may contribute to motivation to perform it well. In summary, for neurodivergent patients, the most informative assessment of likely motion during fMRI is that done during fully informed practise of examples of the language tasks where staying still is a part of the task. We expect that the benefits of training likely also apply to patients without ADHD, consistent with the increased success and lower head motion seen with training across the whole sample.

### Conclusions

This retrospective study utilising clinically acquired task-based language fMRI used for presurgical planning and evaluation in a sample of pediatric epilepsy and neuro-oncology patients, some of whom also had ADHD. We investigated the effects of ADHD, and a tailored language fMRI training program, on success of language lateralisation, and head motion during language fMRI. We found that ADHD was associated with lower language lateralisation success. The training program was associated with higher language lateralization success, particularly in patients with ADHD. ADHD was associated with greater head motion during language fMRI, with some nuanced patterns of results. Although some young patients with ADHD had substantial head motion, most in our sample did not move more than those without ADHD. Training was associated with reduced head motion across all patients. We conclude that the training program increases success of language fMRI and increases data quality in reducing head motion; and that providing individualised support and information can assist patients with ADHD, most of whom are very capable of completing presurgical language fMRI.

## Data Availability

All derived data produced in the present work are illustrated in the manuscript and supplementary materials.

## Acknowledgements

We would like to acknowledge A. Simon Harvey, Amanda Wood (MCRI) and the epilepsy and neuropsychology teams at the Austin Hospital for setting up language fMRI at the RCH; Departments of Child Life Therapy, Neurology, and Neurosurgery at RCH; and the Royal Children’s Hospital Foundation.

