## Supplementary material for "Language fMRI lateralization success and head motion in pediatric epilepsy patients with ADHD, and improvements based on fMRI task training"

### Supplementary information

#### Example of usual script points used in language task training during 15-minute telehealth appointment or as part of Mock fMRI appointment

The exact wording used in task training is different for each patient and family due to tailoring to the age, cognitive and neurodevelopmental abilities of the patient, and in response to their interactions. However, an example of the main points covered and how they may be worded is as follows.

"Thanks for meeting with me. My name is _____, I'm a scientist and I help with the functional MRI scans."
"Have you had an MRI scan here before? How did you find that?"
*[Gauge response to question re any anxiety about scanner or any description of discomfort with noise or small space. Do not directly ask if anxiety as this can prime patients and families to expect anxiety, while most young patients are not innately anxious about going in the scanner]*
"Did you watch a movie or show during that? Do you remember what you watched?"
*[Gauge response re any anxiety or any description of discomfort. This question also allows for building rapport around movie or show choices]*
"This scan is a bit different. Your doctors would like to know where the language parts of your brain are. Those are the parts that do things like listening to words, speaking, and reading. This is so your doctors can understand what the options are for treatment. Does that make sense/do you have any questions?"
*[Gauge whether any questions about purpose of scan]*
"The way can see the language parts of your brain is to have you think of words while you're in the scanner. And when you think of words, we can see the language parts. Does that make sense?"
"The best way we know to do this is to have you do a couple of little language tasks, where you imagine words. I'll show you these now."
*[Show slides with examples of Verb Generation task stimuli]*
"For about ten minutes at the start of the scan, we take a picture of your brain and you just watch a movie. Then I'll talk to you and tell you it's time to do these two tasks. They go for about four minutes each. Then at the end we'll put the movie back on and we'll take any other pictures."
*[Put word 'boat' on the screen]*
"For this first task, we just put a word on the screen, and get you to think of an action word that goes with it. So for this one you might think of 'sailing' or 'swimming'."
"This isn't a test like school, there are no right or wrong answers, we just put something on the screen to help prompt you to imagine words"
*[Put word 'dog' on screen]*
"For this one you might think of 'barking' or 'running'"
*[Put word 'car' on screen]*
"Can you tell me a word that goes with this one?"
*[Gauge whether response given is workable with the parameters of the task:*

- *If, e.g., "driving", say "great" and continue to next example*
- *If, e.g., "you can drive in it", say "That's great! We usually try to think of just one word, like "drive"*
    - *If for multiple following examples, patient provides phrases or sentences rather than verbs, consider that this might reflect their language skills and consider using a version of the task with longer presentation time per stimulus*
- *If, e.g., "it's red", say "Great! Can you think of something you do in a car, like driving?"*
    - *If for multiple following examples, patient provides adjectives or word associations, go ahead with those and encourage patient*
- *If no responses provided and patient is having difficulty with the Verb Generation task, say "No worries! We have another task that you might like better", and continue to the Orthographic Lexical Retrieval task]*

*[Continue with Verb Generation task examples]*
"When we're doing this task, we also stay really nice and still the whole time, for the whole four minutes. You know how if you take a photo of something moving, it comes out blurry? This is just one photo that goes for four whole minutes, and we stay still the whole time so the picture isn't blurry. So make our whole body nice and still and relaxed, keep our face and our arms and legs really relaxed and still. Can you try these next words while staying super nice and still like a statue?
*[Put word example on screen and gauge amount of motion. In young children 7 or 8 years old or developmental delay with presentation approximating this age, if still moving, can say "Now show me how you freeze like a statue! Great! See how long you can stay like a statue! You're doing amazing, what a good statue! Do you think you can have a try of these words while you're a statue? Let's have a go"]*
*[Put another example on screen]*

"Something else we do with this task is just get you to imagine the words in your head, instead of saying them out loud. This is because when we speak, we just naturally move, and we want to stay still so we get a clear picture. Also, when we move our mouth, we can see the parts of your brain that do moving. That's cool but we don't need to see that, we just want to see the thinking of words parts. Let's try some of these but staying nice and still and just imagining the words in your head"
[Put words on screen]
"How was that, did you think of a word"
*[Put rest condition crosshair on screen]*
"For some of the time, there's just a plus sign on the screen. That just means have a rest, you don't need to do anything. Just stay nice and relaxed and still."
"So for this task there's a plus sign on the screen for quite a while, then there are some words, then a plus sign for a while, then some words, and so on, and that goes for four minutes. Do you have any questions about that?"
*[Go on to describe Orthographic Lexical Retrieval task]*
*[Put letter 'A' on the screen]*
"For this task, we just put a letter on the screen and get you to imagine words that start with that letter. Can you tell me out loud any words that start with that one?"
*[Gauge whether responses are workable within the task design:*

- *If, e.g., single word "apple", say "Great! Can you tell me any more?"*
    - *If only one or two examples given, say "No worries if you just think of one or two words! I sometimes can only think of a couple. I can see you were digging around in your mind trying to think of more words, and as long as you do that, that's great! The letter is on the screen for a while, so if you can keep trying to think of more words, that's all we need/*
- *If, .e.g., "Amanda", say, "Great! Can you think of some things, like objects?"*
    - *If patient mostly provides names or place names, say "Great" and continue*
- *If, e.g., "elephant", say "Fantastic! Anything else?"*
- *If patient has difficulty thinking of words/does not provide word responses:*
    - *Can encourage them by saying there are no right or wrong answers, and assure them the letters are just a prompt to help them imagine words*
    - *Consider whether English is a second language and whether examples/responses in first language might be easier*
    - *In older children whose developmental stage might suggest task is possible, consider whether contextual factors may lead to being socially withdrawn, and that patient may be able to do the task on day of scan*
    - *If task appears not possible, discard and say "No worries, thanks for trying that one!"*

*[Optionally and useful for patients with sensory hypersensitivity: "Before and after these word tasks, there will be short, high-pitched beepy-sounding pictures that go for about a minute, that help us get a clear picture"]*
*"So for this scan, there will be ten minutes at the start where we take a structural picture of your brain, and you watch a movie. Then I'll talk to you through your headphones and say it's time to do the word tasks. I'll tell you before each task which one it is, and tell you when it's a good time to have a bit of a stretch and a wriggle if you want. Then after the word tasks we just put the movie back on while we take any other pictures. Does that make sense/do you have any questions? Do you think you'd like some time to wriggle in between the pictures? We can tell you when it's a good time to wriggle your fingers and toes, but it's good to keep your head nice and still. You can tell us if you're comfy or if you need to move or if you want to get out. Does that all sound ok?"*

### Comparison of laterality categories determined based on historical analyses and determined based on reprocessed data

In clinical data processing, preprocessing with fMRIPREP was performed after April 2020 but not before. We reprocessed pre-April 2020 data for standardisation of methods and for consistent estimation of Framewise Displacement. We wanted to ascertain whether differences in processing methods between historical and new data were a factor influencing the feasibility of determining language laterality.  Of the 111 scans prior to April 2020, 101 had reprocessing successfully completed, including preprocessing with fMRIPREP, and processing with FEAT. The output z-statistic activation maps from feat were overlaid on T1 images in lightbox views in axial plane, matching their display for clinical evaluation. Two authors (BA and KS) reviewed these maps and recorded laterality based on visual assessment, which is also the method used in clinical processing. It is important to note that laterality itself was not a measure of interest in the current study, rather the clear presence of a language network in the data without additional presence of other networks that could confound laterality. Of the successfully reprocessed scans, 10 scans had no laterality description historically in EMR for various reasons. For two scans, laterality on reprocessed data did not appear to match with that historically listed in Epic. For one, the description in EMR was ambiguous and referred to superior longitudinal fasciculus (language white matter tract) terminations re possible representation of language. Upon reprocessing with fMRIPREP, this data was classified as left lateralised. For the purpose of the current study, the value on binary categorical variable of 'language laterality determined', was reclassified as determined. For one patient, the description in EMR was bilateral/right, and the reprocessed data showed a right lateralised language network. The laterality results of the reprocessed data do not change treatment for these patients.

**Language lateralisation success based on training, tested separately for first instances and repeat instances of language fMRI**

As described in the main manuscript results section, the factor of 'repeat' could not be explicitly tested due to perfect separation in the data wherein all repeat instances of language fMRI with training had successful language lateralisation. This left the remaining question of solely whether the process of repeating the scan could explain the effect of training. To address this, we ran separate logistic regression models for first instances (n = 212; see Figure X below), and repeat instances (n = 29) of language fMRI.

***Statistical model specification***

For first instances:
glm(lang_laterality_determined ~ training + age_at_scan, data = data_only_first_instances_language_fMRI, family = binomial).

For repeat instances:

Logistic regression in repeat instances (n = 29) of language fMRI failed to converge due to near complete separation, so Firth penalised logistic regression was run:

logistf(formula = lang_laterality_determined ~ training + age_at_scan, data = data_not_first_langfmri)

***Results***

In first instances of language fMRI, logistic regression indicated that training was strongly associated with language lateralisation ($p$ = .003, OR = 5.27, 96%CI 1.92 - 18.6)

In repeat instances of language fMRI, logistic regression indicated there was not strong evidence of training being associated with language lateralisation success ($p$ = 0.078, OR 8.81, 95% CI 0.81–1215), though due to small subsample size, statistical comparisons are not very informative.

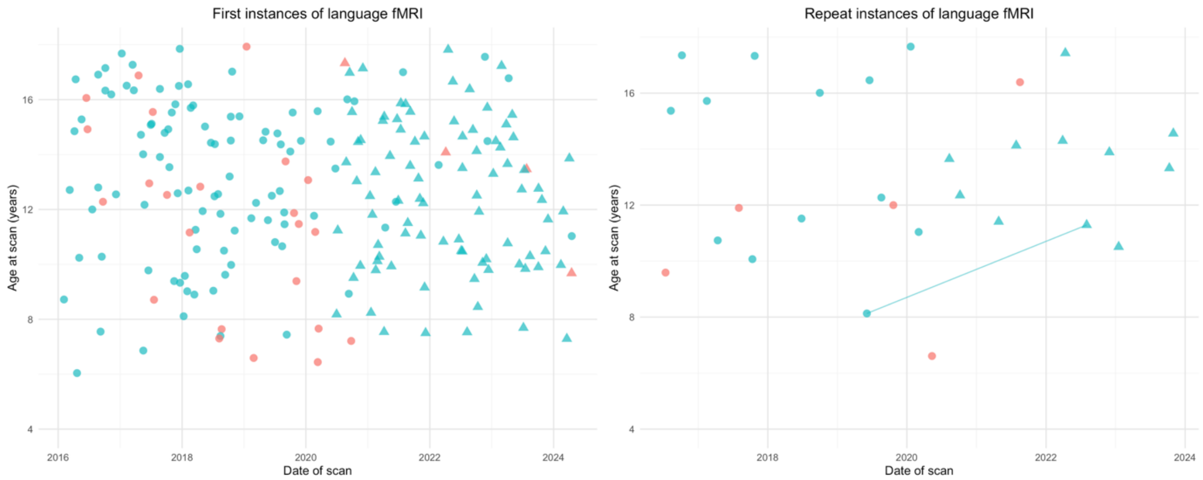


**Figure X.** Success of language lateralization, fMRI training, and age. A) first instance of language fMRI scan. B) repeat instances of language fMRI scans. Green points: language laterality determined, red: language laterality not determined. Triangles: fMRI training provided, circles: fMRI training not provided. Connecting line: repeat scans (2nd and 3rd instances of scan) for individual patient.

#### $FD_{max}$ estimates for CMRR data using TR of 1.5 s rather than 'harmonised' $FD_{max}$

Figure Y shows functional Echo Planar Imaging sequences (Siemens primarily in older acquisitions mostly pre-2018 and CMRR) and Framewise Displacement values based on a) Original TR and b) TR of 3s.

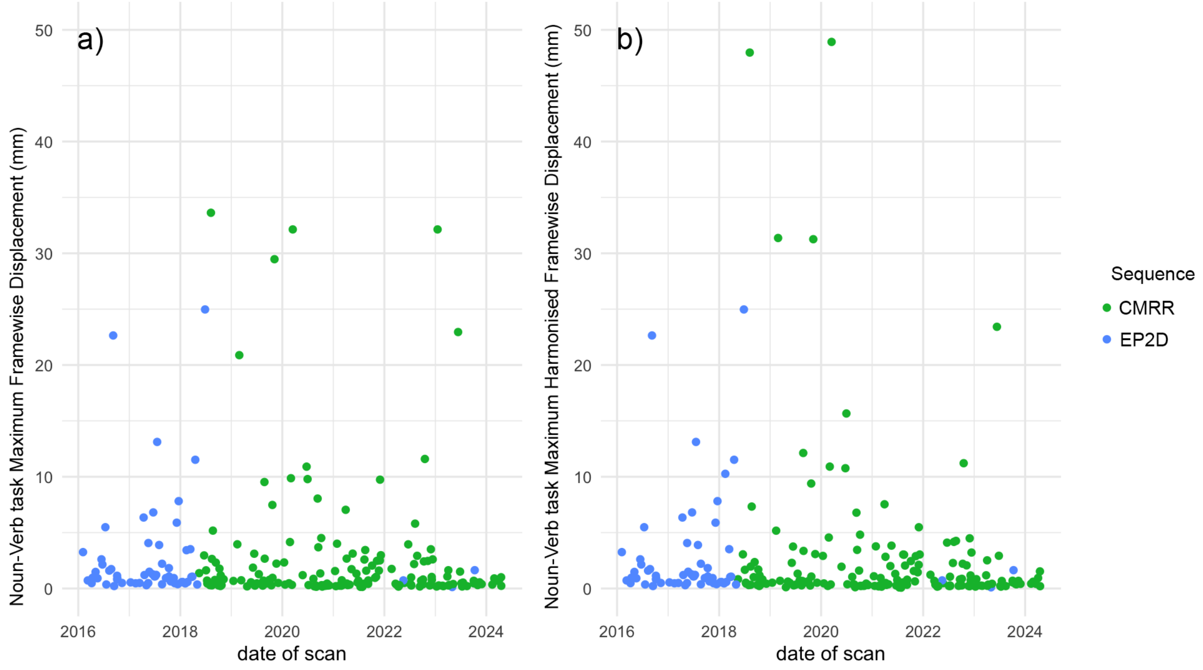


**Figure Y**. Sequence (coded by colour) and estimated Framewise Displacement, shown by date of scan. A) Maximum Framewise displacement for original TR – for Siemens sequences, 1.5 seconds; for CMRR sequences, 3 seconds. B) 'Harmonised' Maximum Framewise displacement where has been calculated for CMRR sequences based on every second volume.

### Head motion using CMRR sequences only, shown separately for patients with and without ADHD and/or autism

We also used the original 1.5 TR FD in a subsample only using CMRR sequences to see whether this effect was still seen with this TR, despite the smaller sample and time range that this allowed. The subset of data with CMRR sequences comprised 150 patients (age 6.59 - 179, *M* 12.5, *SD* 2.72, 86 male, 87 with training provided). 19 had ADHD, 5 had Autism, 4 had both ADHD and Autism.

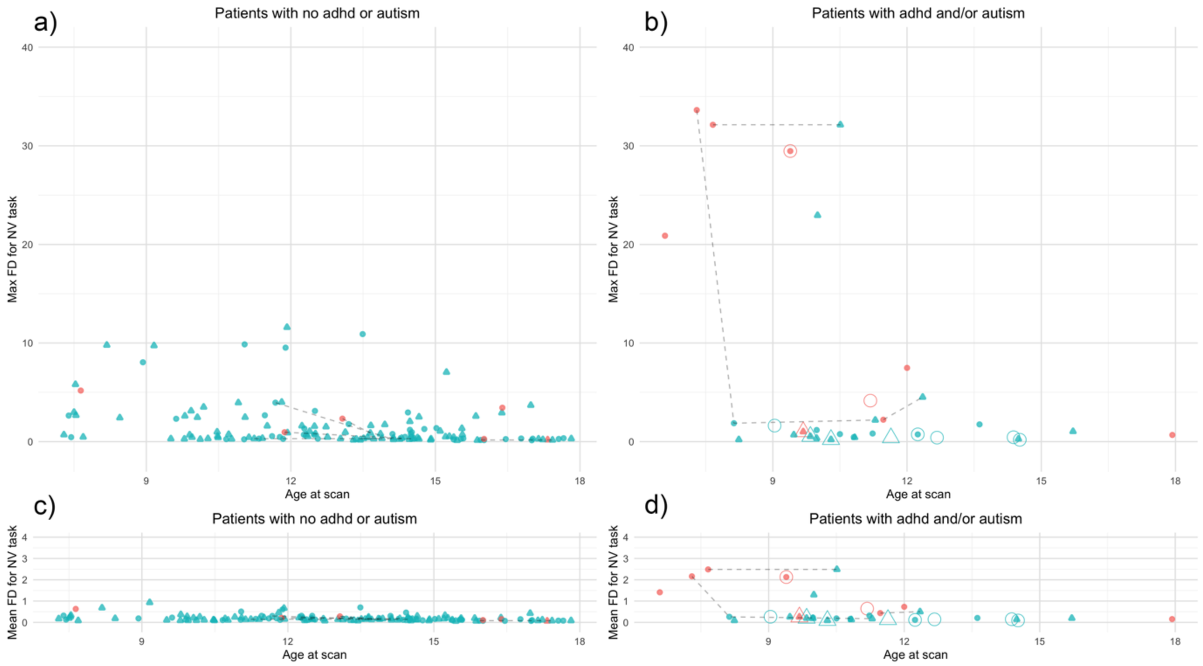


**Figure Z.** Head motion (Framewise Displacement in mm), age, training, and success of language lateralization, for epilepsy patients with diagnoses of ADHD. Green points: language laterality determined, red: language laterality not determined. Triangles: fMRI training provided, circles: fMRI training not provided. B) and d): Small filled points: diagnosis of ADHD, large hollow points: diagnosis of autism. Connecting lines show repeat scans for individual patients.

Linear mixed effects model was run on the subset of data CMRR sequences (i.e., TR = 1.5) with the original non-harmonised maximum framewise displacement metric that uses TR = 1.5, rather than harmonised $FD_{max}$ that used every second volume of the CMRR sequence data to emulate TR = 3. Training, age, and ADHD were specified as fixed effects, and patient ID as a random effect. This model indicated older age was associated with lower $FD_{max}$ ($p < .001$, estimate –0.558, 95%CI -0.8516 - 0.2637), ADHD was associated with higher FDmax ($p < .001$, estimate 4.042, 95%CI 1.810-6.268), training was not found to be significantly associated with FDmax ($p = .113$, estimate –1.264, 95%CI - 2.813-0.285).

Within the subset of data with CMRR sequences, linear mixed effects model was run with the original framewise displacement metric with TR = 1.5, for the subset with ADHD. Age and training were specified as fixed effects, and patient ID as a random effect. This model indicated age was associated with lower $FD_{max}$ ($p = .027$, estimate –1.983, 95%CI -3.612 - -0.3539). Training (provided in 11 of 23 scans) was not significantly associated with $FD_{max}$ ($p = .387$, estimate –3.572, 95%CI -11.426 - 4.1751).
